# Anterolateral temporal lobe localization of dysnomia after temporal lobe epilepsy surgery

**DOI:** 10.1101/2023.09.18.23295718

**Authors:** Asmaa Mhanna, Joel Bruss, Alyssa W. Sullivan, Matthew A. Howard, Daniel Tranel, Aaron D. Boes

## Abstract

**Objectives:** To evaluate what factors influence naming ability after temporal lobectomy in patients with drug-resistant epilepsy.

**Methods:** 85 participants with drug-resistant epilepsy who underwent temporal lobe (TL) resective surgery were retrospectively identified (49 left TL and 36 right TL). Naming ability was assessed before and >3 months post-surgery using the Boston Naming Test (BNT).

Multivariate lesion-symptom mapping was performed to evaluate whether lesion location related to naming deficits. Multiple regression analyses were conducted to examine if other patient characteristics were significantly associated with pre-to post-surgery changes in naming ability.

**Results:** Lesion laterality and location were important predictors of post-surgical naming performance. Naming performance significantly improved after right temporal lobectomy (*p* = 0.015) while a decrement in performance was observed following left temporal lobectomy (*p* = 0.002). Lesion-symptom mapping showed the decline in naming performance was associated with surgical resection of the anterior left middle temporal gyrus (Brodmann area 21, *r* =0.41, *p* = <.001). For left hemisphere surgery, later onset of epilepsy was associated with a greater reduction in post-surgical naming performance (*p* = 0.01).

**Significance:** There is a wide range of variability in outcomes for naming ability after temporal lobectomy, from significant improvements to decrements observed. If future studies support the association of left anterior middle temporal gyrus resection and impaired naming this may help in surgical planning and discussions of prognosis.

## 1. Introduction

Drug-resistant epilepsy constitutes at least 30% of patients with epilepsy (Kwan & Brodie, 2000; Panayiotopoulos, 2007). Epilepsy surgery can be an effective treatment option for many of these patients (Engel et al., 2012; Wiebe et al., 2001). The goal of surgery is to achieve better seizure control while minimizing any adverse effects of undergoing surgery. Temporal lobectomy is the most common resective surgery used for the treatment of intractable seizures in adolescents and adults (Foldvary et al., 2000). It is often effective, with as many as two-thirds of patients being seizure free more than a year after the surgery (Englot & Chang, 2014). However, temporal lobectomy bears a potential risk of cognitive deficits, including, but not limited to the domains of memory, visuospatial ability, and language. A major focus of current research is understanding the neuroanatomical basis of these post-surgical cognitive changes in hopes of informing improvements in surgical planning that reduce the risk of cognitive impairment following surgery without compromising excellent seizure control outcomes.

Language impairment after left temporal lobectomy can include problems with naming, word finding, and verbal fluency. Deficits in naming are especially common, being seen in approximately one third of patients after left temporal lobectomy (Helmstaedter, 2013; Sherman et al., 2011). Difficulty naming can be frustrating for patients and limit one’s ability to communicate clearly in personal and professional settings. It impacts patients’ quality of life, as it can reduce their interest in socially engagement (Farrell et al., 2014). However, the predictors of naming deficits after temporal lobectomy are still unclear and inconsistent across studies.

The left temporal lobe supports key nodes of networks involved in naming. An anatomical framework that has emerged in recent decades includes ventral and anterior temporal cortex supporting multi-modal semantic and conceptual associations that are essential for normal naming performance. Many areas in the left temporal lobe, beyond the classical Broca and Wernicke areas, play an intermediary role in word retrieval. Deficits in picture naming have been linked to lesions of the left anterior and posterior lateral temporal cortex in neurological patients (Bowren Jr et al., 2022; Damasio et al., 1996; Damasio et al., 2004). Using perfusion imaging methods, DeLeon et al. (2007) showed that numerous left hemisphere (LH) areas were crucial for object naming in a group of acute ischemic stroke patients. Deficits were associated with dysfunction of the left anterior temporal lobe, including the superior and middle temporal gyri (DeLeon et al., 2007).

Lesion-symptom mapping is particularly relevant to understanding which brain regions, when surgically resected, are associated with long-term deficits in naming. This approach is clinically important, as many regions with fMRI activity patterns that strongly correlate with naming performance may not be necessary for naming, while a subset of regions will be critical for naming and lesions to those regions will be associated with long-term deficits. In the studies of temporal lobe epilepsy surgery patients reported to date, the critical temporal lobe regions described as being essential for picture naming have been inconsistent across studies. Baldo and colleagues showed that the left mid-posterior middle temporal gyrus and underlying white matter play a critical role in word retrieval associated with an object or picture (Baldo et al., 2013).

Fonseca et al. showed that the posterior temporal areas are essential for naming in temporal lobe epilepsy (Fonseca et al., 2009). More recent studies have shown that the ventral temporal cortex, located mainly in the fusiform gyrus, is significantly associated with naming decline following left temporal lobe epilepsy surgery (Binder et al., 2020; Reindl et al., 2022; Snyder et al., 2023).

In the current study, we aimed to identify the predictors of naming performance after temporal lobe epilepsy surgery in 85 drug-resistant epilepsy patients. Naming was evaluated using the Boston Naming Test (BNT) before surgery and in the chronic epoch after the surgery (>3 months). We hypothesized that there would be a decline in naming after left temporal lobe surgery and that this would be correlated with the resection of critical regions of the left anterior temporal lobe (Bowren Jr et al., 2022; Damasio et al., 1996; Damasio et al., 2004). We applied multivariate lesion-symptom mapping to evaluate whether any areas of the temporal lobe, when resected, are significantly associated with a decline in naming performance. We also aimed to investigate if there are significant associations between other lesion and patient characteristics and postoperative naming performance.

## 2. Methods

### 2.1 Participants

The study sample included 85 participants (45 males, 40 females) who underwent resections in the temporal lobe (49 left TL, 36 right TL) for drug-resistant epilepsy.

Participants were identified from the Patient Registry of the Division of Behavioral Neurology and Cognitive Neuroscience within the Department of Neurology as well as the Department of Neurosurgery at the University of Iowa. The cohort was selected from a pool of 135 patients with medically refractory epilepsy who underwent temporal lobectomy between 1992 and 2021. Patients were included in the analysis if they completed both pre- and post-surgical BNT assessments in the chronic epoch (> 3 months) and had post-surgery brain imaging (MRI or CT scan). Using these criteria 50 patients were excluded. One patient did not have post-surgery brain imaging, and 49 did not have both pre- and post-surgery BNT scores. Our institutional ethical review committee approved of the study design in advance of data collection and analysis. The demographic characteristics of the sample are presented in Table 1.

**Table 1.** Study population characteristics.

|  | **Left temporal lobectomy**<br>**(N = 49)** | **Right temporal lobectomy**<br>**(N = 36)** |
| --- | --- | --- |
| Sex | 27 females (55.1%) | 18 females (50%) |
| Handedness | 43 Right-handed (87.7%)<br>4 Left-handed (8.2%)<br>2 Ambidextrous (4.1%) | 30 Right-handed (83.3%)<br>4 Left-handed (11.1%)<br>2 Ambidextrous (5.6%) |
| Language dominance (based on WADA test or fMRI) | 38 Left (77.5%)<br>4 Right (8.2%)<br>3 Mixed (6.1%)<br>4 Unknown (8.2%) | 25 Left (69.4%)<br>3 Right (8.3%)<br>2 Mixed (5.6%)<br>6 Unknown (16.7%) |
| Seizure onset age (range; mean ± SD; median) | 7 mo. – 59 yrs.;<br>17.017 ± 14.61 yrs.;<br>17.5 yrs. | 6 mo. – 38 yrs.;<br>15.76 ± 11.51 yrs.;<br>14 yrs. |
| Age at surgery (range; mean ± SD; median) | 17.8 – 62.3 yrs.;<br>39.20 ± 12.22 yrs.;<br>37.4 yrs. | 17.5 – 59 yrs.;<br>34.01 ± 11.54 yrs.;<br>32.7 yrs. |
| Seizure freedom after surgery | 34 (69.4%) seizure-free/<br>Engel class 1.<br>15 (30.6%) Engel class 2,3,<br>or 4. | 31 (86.1%) seizure-free/<br>Engel class 1.<br>5 (13.9%) Engel class 2,3, or 4. |
| BNT* post-surgical assessment time (range; mean ± SD; median) | 3.5 mo. – 4.26 yrs.;<br>1.05 ± 0.87 yrs.;<br>9 mo. | 3 mo. – 9.24 yrs.;<br>1.90 ± 2.09 yrs.;<br>1.21 yrs. |
| Education (range; mean ± SD; median) | 8 – 18 yrs.;<br>13.06 ± 2.22 yrs.;<br>12 yrs. | 11 – 18 yrs.;<br>14.03 ± 2.02 yrs.;<br>14 yrs. |
| *BNT = Boston Naming Test |  |  |

### 2.2 Picture naming test

Picture naming was assessed using the standard 60-item version of the BNT before surgery and at least 3 months following surgery. The BNT is a measure of confrontational word retrieval, classically used to evaluate patients with aphasia (Goodglass et al., 1983). It consists of 60-line drawings of objects, graded in difficulty. An item is considered passed if the patient either spontaneously names the item correctly within 20 seconds or names the item correctly within 20 seconds after being given a stimulus cue from the examiner. The assessments were performed following the standard testing and scoring procedure. The BNT change score was computed by subtracting the preoperative from the postoperative score. We also calculated the percent change score using the following equation: (100*(postoperative score – preoperative score) / preoperative score).

### 2.3 Brain imaging and lesion segmentation

All participants underwent brain MRI or CT scanning after surgery. For patients who registered in the Patient Registry prior to 2006, the surgical resection cavity was manually drawn for each participant using the MAP-3 lesion tracing method, which entails manually tracing lesion borders on a template brain (Damasio & Frank, 1992; Fiez et al., 2000). Following 2006, lesions were manually drawn on native T1-weighted or CT images (Smith et al., 2004) and then converted to the 1-mm MNI152 template brain using a high-deformation, non-linear, enantiomorphic, registration procedure from the Advanced Normalization Tools (Avants et al., 2008; Brett et al., 2001; Nachev et al., 2008). The anatomical accuracy of the lesion boundaries was reviewed and edited as needed in native and MNI space by a neurologist (A.D.B.) blinded to naming outcomes.

### 2.4 Statistical analysis and lesion-symptom mapping

We applied multivariate lesion-symptom mapping to the behavioral and lesion data across all 85 patients to identify brain structures that, when resected, are significantly associated with changes in BNT. We used the LESYMAP package in R (https://github.com/dorianps/LESYMAP), which uses sparse canonical correlation analysis for neuroimaging (SCCAN) (Pustina et al., 2018). The SCCAN method involves an optimization procedure to assign a continuous weight to each voxel (range 0 -1) in order to maximize the multivariate correlation with the true behavioral scores. The validity of the map is evaluated using a 4-fold cross-validation (with 25% of the sample held out at each fold). The optimal sparseness and statistical significance of the lesion-symptom association is tested by comparing the actual change in naming ability to that predicted by the model. Coordinates of significant clusters were presented in the Montreal Neurological Institute (MNI) space and displayed on the MNI152 standard-space T1-weighted average structural template image for 3D visualization in Slicer software.

We also evaluated lesion intersection with other sites previously identified in association with left temporal lobectomy postoperative dysnomia. For this analysis, we produced a 1 cm spherical region of interest centered at the peak site in lesion-symptom mapping studies of impaired naming after temporal lobe epilepsy surgery from (Baldo et al., 2013; Binder et al., 2020; Reindl et al., 2022; Snyder et al., 2023). For each ROI we divided our cohort into those with lesion masks that intersected or did not intersect with these ROIs and compared the post-surgical naming outcome between the two groups.

Statistics of other demographic and behavioral data were performed using IBM SPSS Statistics 20.0 (http://www.spss.com). Statistical analysis of differences between pre- and post-surgical naming scores within each group was performed using paired t-tests. Stepwise multiple linear regression analyses were conducted to identify patient and lesion characteristics that significantly associated with pre to post-surgery changes in naming ability.

## 3 Results

### 3.1 Behavioral performance findings

The overall naming performance did not significantly change from pre-to post-surgery when all 85 participants were included together (Table 2). When divided into groups based on hemisphere, individuals with right TL surgery had a significant improvement in the BNT score postoperatively, with 64% of patients having a higher postoperative score compared to the baseline performance. As a group, participants with right hemisphere surgery showed BNT score improvement by an average of 1.7 points (SD = 3.9, *p* = 0.015; or 4.26% (SD = 10.56%) with a broad range (-16.6 – 50). For the left TL group, there was a significant decline in naming performance after surgery, with 57.14% showing lower postoperative score compared to baseline performance. They had an average decline of 3.43 points (SD = 7.4, *p* = 0.002; or 6.04% (SD = 17.27%). 36.73% showing a decline of 10% or more in their naming score, and 14.29% showing a decline of 20% or more. Six patients (12.24%) showed decline of more than 30%.

**Table 2.** BNT pre-to post-surgery change scores in right and left temporal lobectomy groups.

| Left Temporal Lobectomy |  |  |  | Right Temporal Lobectomy |  |  |  |
| --- | --- | --- | --- | --- | --- | --- | --- |
| ID | Preoperative BNT score | Postoperative BNT score | BNT change | ID | Preoperative BNT score | Postoperative BNT score | BNT change |
| 1 | 43 | 44 | 1 | 1 | 40 | 49 | 9 |
| 2 | 51 | 45 | -6 | 2 | 52 | 55 | 3 |
| 3 | 52 | 47 | -5 | 3 | 54 | 58 | 4 |
| 4 | 40 | 40 | 0 | 4 | 55 | 58 | 3 |
| 5 | 40 | 21 | -19 | 5 | 57 | 51 | -6 |
| 6 | 55 | 48 | -7 | 6 | 47 | 51 | 4 |
| 7 | 46 | 41 | -5 | 7 | 54 | 58 | 4 |
| 8 | 58 | 57 | -1 | 8 | 53 | 55 | 2 |
| 9 | 36 | 42 | 6 | 9 | 59 | 60 | 1 |
| 10 | 53 | 57 | 4 | 10 | 53 | 52 | -1 |
| 11 | 31 | 34 | 3 | 11 | 50 | 53 | 3 |
| 12 | 46 | 46 | 0 | 12 | 54 | 55 | 1 |
| 13 | 29 | 28 | -1 | 13 | 49 | 53 | 4 |
| 14 | 19 | 25 | 6 | 14 | 55 | 55 | 0 |
| 15 | 33 | 20 | -13 | 15 | 56 | 55 | -1 |
| 16 | 47 | 42 | -5 | 16 | 60 | 60 | 0 |
| 17 | 55 | 53 | -2 | 17 | 60 | 60 | 0 |
| 18 | 59 | 36 | -23 | 18 | 49 | 51 | 2 |
| 19 | 57 | 47 | -10 | 19 | 51 | 53 | 2 |
| 20 | 56 | 48 | -8 | 20 | 56 | 57 | 1 |
| 21 | 44 | 41 | -3 | 21 | 26 | 39 | 13 |
| 22 | 55 | 51 | -4 | 22 | 53 | 49 | -4 |
| 23 | 50 | 43 | -7 | 23 | 57 | 57 | 0 |
| 24 | 43 | 53 | 10 | 24 | 56 | 56 | 0 |
| **25** | 43 | 51 | 8 | **25** | 49 | 52 | 3 |
| **26** | 55 | 48 | -7 | **26** | 55 | 59 | 4 |
| **27** | 58 | 55 | -3 | **27** | 54 | 45 | -9 |
| **28** | 57 | 59 | 2 | **28** | 53 | 57 | 4 |
| **29** | 11 | 14 | 3 | **29** | 58 | 60 | 2 |
| **30** | 56 | 51 | -5 | **30** | 40 | 47 | 7 |
| **31** | 54 | 55 | 1 | **31** | 55 | 55 | 0 |
| **32** | 48 | 49 | 1 | **32** | 41 | 45 | 4 |
| **33** | 45 | 50 | 5 | **33** | 44 | 47 | 3 |
| **34** | 54 | 55 | 1 | **34** | 58 | 58 | 0 |
| **35** | 46 | 41 | -5 | **35** | 45 | 41 | -4 |
| **36** | 46 | 44 | -2 | **36** | 50 | 52 | 2 |
| **37** | 36 | 27 | -9 |  |  |  |  |
| **38** | 56 | 57 | 1 |  |  |  |  |
| **39** | 48 | 51 | 3 |  |  |  |  |
| **40** | 45 | 45 | 0 |  |  |  |  |
| **41** | 53 | 37 | -16 |  |  |  |  |
| **42** | 55 | 59 | 4 |  |  |  |  |
| **43** | 49 | 51 | 2 |  |  |  |  |
| **44** | 56 | 46 | -10 |  |  |  |  |
| **45** | 54 | 45 | -9 |  |  |  |  |
| **46** | 55 | 37 | -18 |  |  |  |  |
| **47** | 43 | 21 | -22 |  |  |  |  |
| **48** | 52 | 53 | 1 |  |  |  |  |
| **49** | 57 | 52 | -1 |  |  |  |  |

### 3.2 Lesion analysis findings

Lesion overlap across all patients is shown in Fig 1a. The lesion-symptom mapping analysis revealed the strongest correlation of BNT decline and resection of the anterior left middle temporal gyrus (Brodmann area 21), with a peak voxel at MNI coordinate (-58, 2, -27) (*r* =0.41, *p* = <.001 Figure 1b). Lesion intersection with this peak coordinate was associated with reduced naming performance relative to individuals with left hemisphere surgery that did not overlap with this site (mean -3.78, SD = 6.39; t = 3.57, *p* < 0.001). There was a significant difference in pre-to post-surgery naming scores across all the BNT subcategories (animals, plants, and tools) among patients with overall BNT decline after surgery and lesion masks intersecting with the peak coordinate (see Table 1 in the supplementary). A smaller and less robust second cluster was observed on the anteromedial temporal cortex partially overlapping the amygdala (MNI -29, 0, -24).

**Fig. 1.**
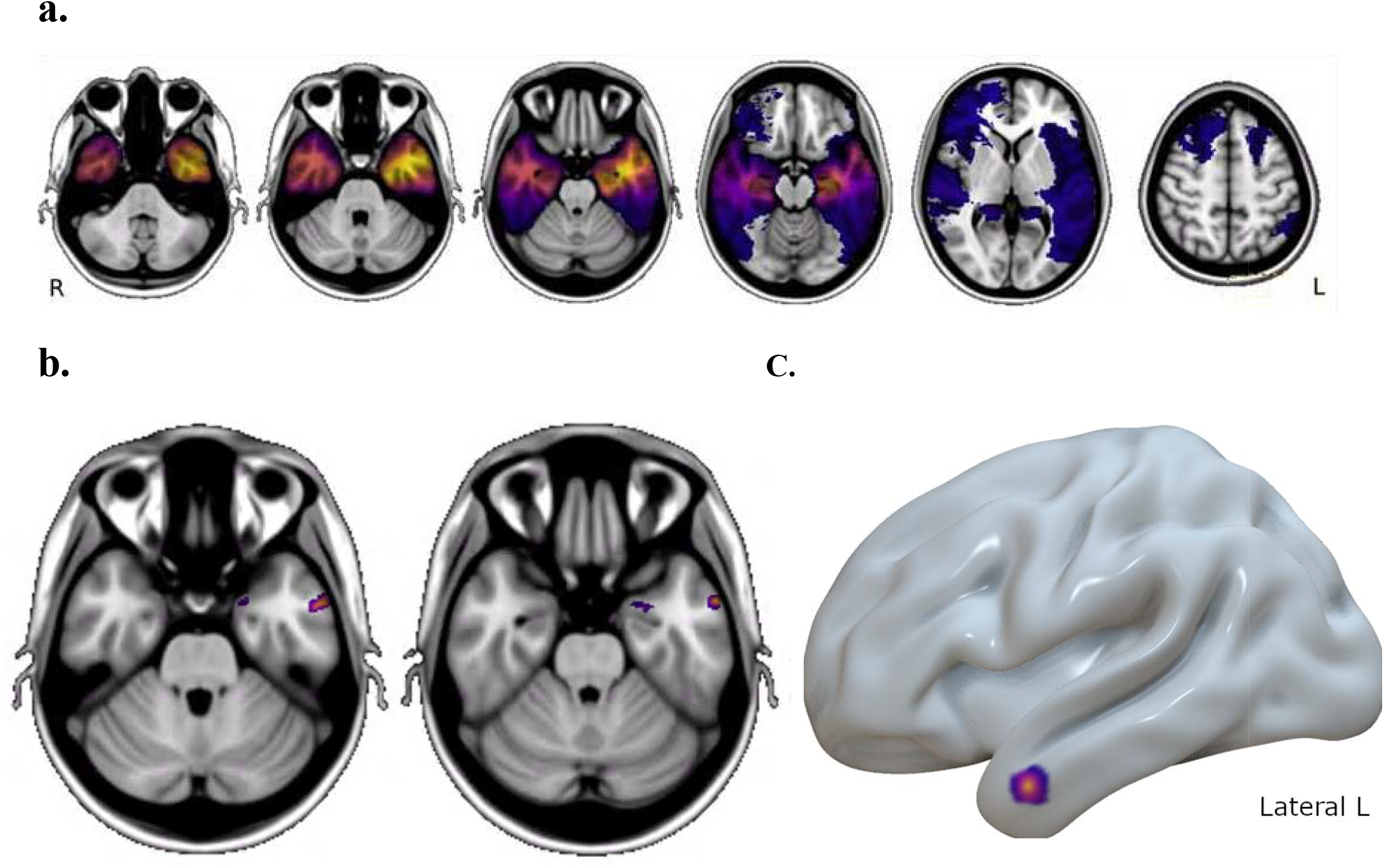
a. Lesion overlap. Lesion overlap map of the surgical resection cavities used in this analysis, **b, c Multivariate lesion symptom mapping**. The lesion-symptom map of participants with left hemisphere lesions, showing an association of the anterior third of the left middle temporal gyrus resections with decline in post-surgical naming ability (r =0.41, *p* <.001.

Lesion intersection with other peak sites associated with dysnomia from previously published studies was also tested to evaluate naming outcome between subjects with resections that overlap with these peak regions and those that did not. Individuals with lesions intersecting with these sites showed greater naming decline than those who did not (Fig. 2). These differences did not reach statistical significance, likely due to low statistical power.

**Fig. 2.**
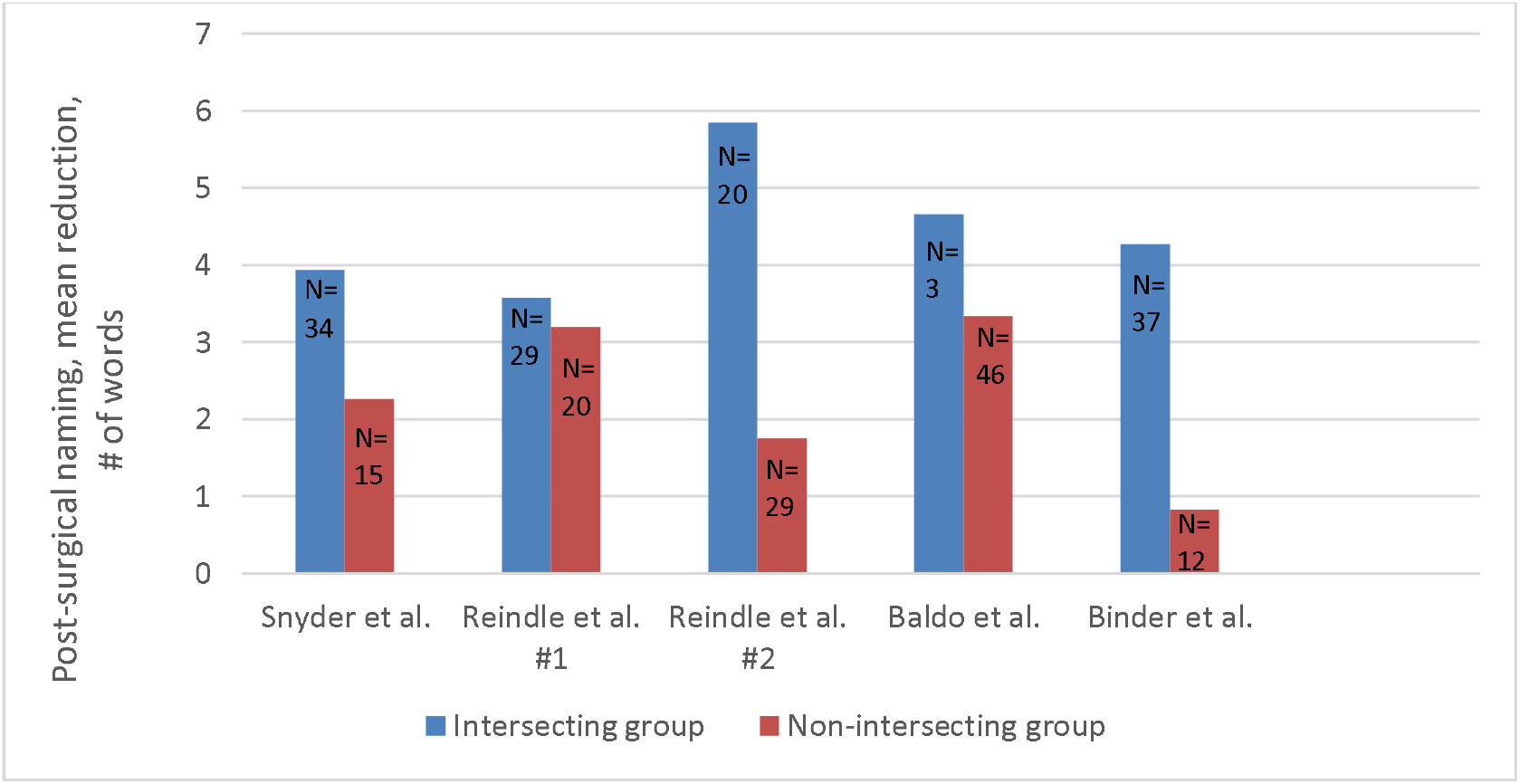
Naming outcome prediction after left TL surgery based on lesion intersection with other peak sites associated with dysnomia from published studies. Individuals with lesions intersecting with these sites showed greater naming decline after surgery than those who did not, though these differences did not reach statistical significance. MNI coordinates for regional peaks include: Snyder et al. (MNI -33.1, -12.3, -33.4), Reindle et al. #1 (MNI -52, -12, -37), Reindle et al. #2 (MNI -38, -21, -32), Baldo et al. (MNI -46, -48, 0) and Binder et al. (-35, -25, -19).

### 3.3 Other variables associated with naming outcomes

For both right and left TL groups, there was no significant association between naming change after surgery and age at surgery, duration of epilepsy, time of naming assessment after surgery, seizure freedom after surgery, lesion volume, or years of education. When analyzed by hemisphere, the RH group with higher preoperative naming performance showed a greater decline postoperatively (β = -0.42, *p* < 0.001). For each point of increase in preoperative naming score, there would be a 0.42-point decline in the naming score after right TL. For the left hemisphere group, patients with postoperative deterioration of naming performance were significantly older at the time of epilepsy onset (β = -0.42, *p* < 0.001).

## 4. Discussion

This study investigated the predictors of naming performance after temporal lobe surgery in patients with drug-resistant epilepsy. We identified a region of the left anterior middle temporal gyrus (Brodmann area 21), that, when resected, was significantly associated with a decline in naming performance. Our findings showed that lesion laterality and location were important predictors of post-surgical naming performance. Those with left hemisphere surgery showed impaired post-surgical naming as a group, this was especially the case for patients with later onset of epilepsy. In contrast, significant improvement in naming performance was observed in the group with right temporal lobe surgery. For the purposes of this discussion, we will focus on two main findings: 1) the anatomical findings of our lesion-symptom map, and 2) the observation of improved performance following right temporal lobectomy.

Our results build upon earlier findings showing postoperative picture naming impairments associated with left temporal lobe surgery (Fonseca et al., 2009; Hamberger, 2015; Hermann et al., 1999; Langfitt & Rausch, 1996; Schwarz et al., 2005). These anatomical results contribute to a growing body of research performing lesion-symptom mapping of epilepsy surgery (Binder et al., 2020; Reindl et al., 2022; Snyder et al., 2023). These studies have highlighted critical anatomical nodes of more posterior temporal locations along the fusiform gyrus. In our sample, the strongest anatomical association with dysnomia was with lesions of the anterolateral middle temporal gyrus. This region has not been reported in prior temporal lobectomy lesion-symptom mapping studies, but this region has been implicated in naming in prior lesion and functional imaging work (Bowren Jr et al., 2022; Breining et al., 2023; Damasio et al., 1996; Damasio et al., 2004; DeLeon et al., 2007). In fact, the significant finding displayed in Fig. 1c overlaps with a prior lesion-symptom map of naming performance in an independent (non-overlapping) cohort of 432 individuals published by our group (Bowren Jr et al., 2022). These different regions of the left temporal lobe language networks (anterior middle temporal gyrus versus fusiform gyrus) may contribute differentially to naming, with prior work implicating the anterior temporal cortex more in semantic processing (Baldo et al., 2013; Damasio et al., 2004; Fonseca et al., 2009; Price et al., 2005; Raymer et al., 1997).

While anatomy of the resection cavity is an important factor in naming outcomes, it likely interacts with other factors. We observed that later onset of epilepsy was associated with a greater reduction in naming ability relative to individuals with earlier onset of epilepsy. This is in line with previous research (Bell et al., 2002; Davies et al., 2005; Hermann et al., 1995; Ruff et al., 2007; Saykin et al., 1995; Yucus & Tranel, 2007). Stafiniak and colleagues found that age of onset of epilepsy after age 5 was associated with a significant decline (≥25%) in naming after left TL resection compared to subjects with an earlier age of onset of epilepsy (≤5 years) (Stafiniak et al., 1990). In addition, Reindl and colleagues showed deterioration in naming performance six months after dominant TL surgery in patients with epilepsy onset after 5 years of age but not with earlier epilepsy onset (Reindl et al., 2022). This may be attributed to brain plasticity in patients who experience epileptic seizures at an early age (Chou et al., 2018), which could facilitate language reorganization and thus greater resilience to surgical resection of the seizure focus (Bell et al., 2002; Devinsky et al., 1993; Ruff et al., 2007; Saykin et al., 1995).

The second main finding we wish to highlight is the improvement in naming performance observed after right temporal lobe epilepsy surgery. To the best of our knowledge, this was the first study that showed significant improvement in naming performance after right temporal lobectomy, possibly because naming is less commonly studied in association with right hemisphere surgery. This observed improvement may be secondary to overall improvement in epilepsy, with fewer epileptiform discharges, less frequent seizures, or fewer side effects of seizure medications. A few studies investigated the effect of right TL on visual naming that did not show a change in BNT performance (Escorsi-Rosset et al., 2011) or naming of living and non-living entities (Tippett et al., 1996).

We did not find any significant association between naming change after surgery and age at surgery, duration of epilepsy, time of naming assessment after surgery, seizure freedom after surgery, lesion volume, or years of education for either the right TL or left TL groups. These factors have been variably associated with naming outcomes in prior studies. However, the most consistent predictors across most studies are age at onset of epilepsy and baseline BNT scores (Reindl et al., 2022; Schwarz et al., 2005).

Our study has limitations. The study included a wide range of patients in terms of onset of epilepsy, age at the time of surgery, and the variable timing after surgery for assessing naming, ranging from 3 months up to 9 years. Notably we did not observe a significant effect of time since surgery on naming performance, suggesting that performance largely stabilizes after 3 months. We focused on the BNT and did not include other categories of visual naming (e.g., famous faces, landmarks) or other modalities (e.g., auditory naming). Future studies could investigate lesion-symptom mapping for more specific categories of picture naming as well as for auditory naming. In addition, our study did not distinguish if the naming deficit after left TL is due to a concept identification (semantic) deficit or a word retrieval (lexical) deficit.

In summary, our study highlighted the role of lesion laterality and location in postoperative naming outcomes following temporal lobe epilepsy surgery. We identified the left anterior middle temporal gyrus as a critical region for naming in drug-resistant epilepsy patients. We also showed that postoperative naming improved, on average, amongst individuals with a right temporal lobectomy. If future studies support the association of anterior left middle temporal gyrus resection and impaired naming this may help in surgical planning and discussions of prognosis.

## Author Contributions

A.M. conducted the data analysis and drafted the manuscript and figures.

J.B. contributed to data collection, data analyses, and figures. A.W.S., M.A.H. and D.T. contributed to the data acquisition and the revision of the manuscript. A.D.B. contributed to the conceptualization, design, and supervision of the study. All authors revised and approved the final version of the manuscript.

## Data Availability

The datasets generated and/or analyzed during the current study are available upon request from the corresponding author.

## Ethics approval

All procedures performed in the study were in accordance with the University of Iowa Institutional Review Board.

## Consent to participate

Informed consents were obtained from all participants in accordance with the University of Iowa Institutional Review Board.

## Consent to publish

Not applicable

