## Supplemental Table 1 for "Anterolateral temporal lobe localization of dysnomia after temporal lobe epilepsy surgery"

|  | Page |
| --- | --- |
| Title | 1 |
| Supplementary Table 1. BNT pre- to post-surgery change scores across different subcategories among patients with overall BNT decline after surgery and lesion masks intersecting with our peak coordinate. | 2 |

| **ID** | **Animals BNT scores pre-TL** | **Animals BNT scores post-TL** | **Animals BNT change scores** | **Plants BNT scores pre-TL** | **Plants BNT score post-TL** | **Plants BNT change scores** | **Tools BNT scores pre-TL** | **Tools BNT scores post-TL** | **Tools BNT change scores** | **Total- BNT scores pre-TL** | **Total BNT scores post-TL** | **Total BNT Change scores** |
| --- | --- | --- | --- | --- | --- | --- | --- | --- | --- | --- | --- | --- |
| **1** | 7 | 6 | -1 | 5 | 5 | 0 | 39 | 34 | -5 | 51 | 45 | -6 |
| **2** | 8 | 7 | -1 | 5 | 5 | 0 | 39 | 35 | -4 | 52 | 47 | -5 |
| **3** | 1 | 1 | 0 | 5 | 3 | -2 | 33 | 17 | -16 | 40 | 21 | -19 |
| **4** | 8 | 7 | -1 | 6 | 6 | 0 | 41 | 35 | -6 | 55 | 48 | -7 |
| **5** | 5 | 5 | 0 | 6 | 5 | -1 | 35 | 31 | -4 | 46 | 41 | -5 |
| **6** | 8 | 8 | 0 | 6 | 6 | 0 | 44 | 43 | -1 | 58 | 57 | -1 |
| **7** | 5 | 5 | 0 | 4 | 4 | 0 | 20 | 19 | -1 | 29 | 28 | -1 |
| **8** | 5 | 1 | -4 | 4 | 3 | -1 | 24 | 16 | -8 | 33 | 20 | -13 |
| **9** | 7 | 5 | -2 | 5 | 5 | 0 | 35 | 32 | -3 | 47 | 42 | -5 |
| **10** | 7 | 8 | 1 | 6 | 5 | -1 | 42 | 40 | -2 | 55 | 53 | -2 |
| **11** | 8 | 5 | -3 | 6 | 4 | -2 | 45 | 27 | -18 | 59 | 36 | -23 |
| **12** | 8 | 8 | 0 | 5 | 5 | 0 | 43 | 35 | -8 | 56 | 48 | -8 |
| **13** | 6 | 6 | 0 | 5 | 5 | 0 | 33 | 30 | -3 | 44 | 41 | -3 |
| **14** | 8 | 8 | 0 | 6 | 6 | 0 | 41 | 37 | -4 | 55 | 51 | -4 |
| **15** | 8 | 8 | 0 | 6 | 5 | -1 | 36 | 30 | -6 | 50 | 43 | -7 |
| **16** | 8 | 7 | -1 | 6 | 6 | 0 | 41 | 35 | -6 | 55 | 48 | -7 |
| **17** | 8 | 8 | 0 | 6 | 6 | 0 | 44 | 41 | -3 | 58 | 55 | -3 |
| **18** | 8 | 7 | -1 | 6 | 6 | 0 | 42 | 38 | -4 | 56 | 51 | -5 |
| **19** | 7 | 7 | 0 | 6 | 4 | -2 | 33 | 30 | -3 | 46 | 41 | -5 |
| **20** | 8 | 7 | -1 | 5 | 5 | 0 | 33 | 32 | -1 | 46 | 44 | -2 |
| **21** | 4 | 3 | -1 | 6 | 4 | -2 | 26 | 20 | -6 | 36 | 27 | -9 |
| **22** | 8 | 6 | -2 | 6 | 4 | -2 | 39 | 27 | -12 | 53 | 37 | -16 |

Supplementary Table 1. BNT pre- to post-surgery change scores across different subcategories (animals, plants, and tools) among patients with overall BNT decline after surgery and lesion masks intersecting with our peak coordinate.
